# Differential Effects of Intervention Timing on COVID-19 Spread in the United States

**DOI:** 10.1101/2020.05.15.20103655

**Authors:** Sen Pei, Sasikiran Kandula, Jeffrey Shaman

## Abstract

Assessing the effects of early non-pharmaceutical interventions^1-5^ on COVID-19 spread in the United States is crucial for understanding and planning future control measures to combat the ongoing pandemic^6-10^. Here we use county-level observations of reported infections and deaths^11^, in conjunction with human mobility data^12^ and a metapopulation transmission model^13,14^, to quantify changes of disease transmission rates in US counties from March 15, 2020 to May 3, 2020. We find significant reductions of the basic reproductive numbers in major metropolitan areas in association with social distancing and other control measures. Counterfactual simulations indicate that, had these same control measures been implemented just 1-2 weeks earlier, a substantial number of cases and deaths could have been averted. Specifically, nationwide, 56.5% [95% CI: 48.1%-65.9%] of reported infections and 54.0% [95% CI: 43.6%-63.8%] of reported deaths as of May 3, 2020 could have been avoided if the same control measures had been implemented just one week earlier. We also examine the effects of delays in re-implementing social distancing following a relaxation of control measures. A longer response time results in a stronger rebound of infections and death. Our findings underscore the importance of early intervention and aggressive response in controlling the COVID-19 pandemic.

## Main text

The ongoing COVID-19 pandemic has caused millions of infections and hundreds of thousands of deaths worldwide^8,9^. In the United States, the first imported case of COVID-19 was reported on January 20, 2020^10^. In subsequent weeks, community transmission of COVID-19 was established, and the causative pathogen, SARS-CoV-2, quickly spread throughout the entire country^9^. As of May 14, 2020, over 1.4 million infections and 84 thousand deaths had been confirmed nationwide^11^, making the US the hardest-hit country in the world to date.

In an effort to slow the spread of COVID-19, control measures enforcing social distancing and restricting individual contact were implemented across the US beginning in mid-March. In other countries, these non-pharmaceutical interventions (NPIs) have successfully controlled the spread of COVID-19^1-5^; however, in the US, the effectiveness of these control measures has been less pronounced. It is therefore important that changes in virus transmissibility within the US, due to NPIs, be quantified, so that the effects of earlier interventions on cases and deaths can be evaluated.

In this study we adapted and applied a dynamic metapopulation model informed by human mobility data and representing SARS-CoV-2 transmission in 3142 US counties (Methods). We explicitly simulate documented and undocumented infections^13^, for which separate transmission rates, *β* and *μβ* (*μ* < 1), respectively, are defined. Here *μ* is the relative transmissibility of undocumented infections. To reflect heterogeneity in transmission rates across the US while avoiding a large number of model parameters, we defined a separate *β_i_* for counties with greater than 400 cumulative confirmed cases as of May 3, 2020 (n=311). The remaining 2831 counties were apportioned among 16 additional transmission rate parameters depending on cumulative case levels and population density (Methods). Other parameters in the model include the ascertainment rate, *α*, which represents the fraction of infections documented as confirmed cases, the average latency period, *Z*, the average duration of infection, *D*, and the travel multiplicative factor, *θ*.

Parameter estimation was performed using the ensemble adjustment Kalman filter (EAKF)^15^ in conjunction with county-level observations of both daily reported cases and deaths in the US from February 21, 2020 to May 3, 2020 (Methods). As the model parameters may not be as well constrained in counties with low case counts, in this study, we focus on several metropolitan areas with large populations and abundant data (Methods), for which parameter estimates are better informed.

Daily cases and deaths in the US and the New York metropolitan area are well fit by the transmission model (Fig.1 a-d). The inferred basic reproductive numbers, *R*_0_ *= βD[α +*(1 − *α*)*μ*]^13,16^, for six metropolitan areas – New York, New Orleans, Los Angeles, Chicago, Boston and Miami – on five dates (March 15, March 29, April 12, April 26, May 3) are shown in Table 1 (Methods). After March 15, *R*_0_ in all six metropolitan areas decreases substantially in association with the implementation of social-distancing policies and practices. The estimated effective reproductive numbers, *R_e_ ≡ βD[α +(*1 − *α)μ]S/N*, for these six metropolitan areas also decrease from March 15, 2020 to May 3, 2020 (Fig. 1e). In four of the six metropolitan areas *R_e_* is well below 1 as of May 3, 2020. For Chicago and Los Angeles, where daily confirmed cases and deaths are still increasing or stable, *R_e_* is close to 1 (Figs. S3-S4). In the New York metropolitan area *R_e_* dropped below 1 on April 8 and has continued decreasing since then. The estimated nationwide ascertainment rate declined from 0.18 on March 15, a time of rapid COVID-19 spread, and went below 0.1 on March 30 (Fig. 1f). The ascertainment rate slowly increased after April 5. Note that this finding indicates that, prior to April 5, even though testing capacity increased substantially, daily new infections increased faster, leading to a declining ascertainment rate.

**Table 1.** Estimated basic reproductive numbers *(R*_0_) for the New York, New Orleans, Los Angeles, Chicago, Boston and Miami metropolitan areas on March 15, March 29, April 12, April 26 and May 3. Mean estimate (95% CIs) are presented.

|  | March 15 | March 29 | April 12 | April 26 | May 3 |
| --- | --- | --- | --- | --- | --- |
| New York | 2.82<br>(2.53, 3.07) | 1.71<br>(1.60, 1.88) | 0.83<br>(0.72, 0.97) | 0.53<br>(0.44, 0.63) | 0.42<br>(0.34, 0.50) |
| New Orleans | 2.42<br>(2.05, 2.82) | 1.83<br>(1.58, 2.09) | 0.63<br>(0.49, 0.76) | 0.50<br>(0.38, 0.62) | 0.44<br>(0.33, 0.54) |
| Los Angeles | 2.88<br>(2.33, 3.53) | 1.50<br>(1.26, 1.80) | 1.16<br>(0.98, 1.37) | 0.84<br>(0.66, 1.07) | 0.78<br>(0.57, 1.00) |
| Chicago | 2.75<br>(2.42, 3.07) | 1.90<br>(1.69, 2.14) | 1.15<br>(1.00, 1.33) | 1.10<br>(0.91, 1.27) | 1.24<br>(0.94, 1.45) |
| Boston | 3.18<br>(2.57, 3.79) | 2.05<br>(1.77, 2.32) | 1.39<br>(1.19, 1.56) | 0.38<br>(0.32, 0.52) | 0.16<br>(0.13, 0.21) |
| Miami | 2.62<br>(2.21, 3.14) | 1.45<br>(1.25, 1.75) | 0.96<br>(0.84, 1.15) | 0.51<br>(0.40, 0.66) | 0.34<br>(0.26, 0.45) |

**Figure 1.**
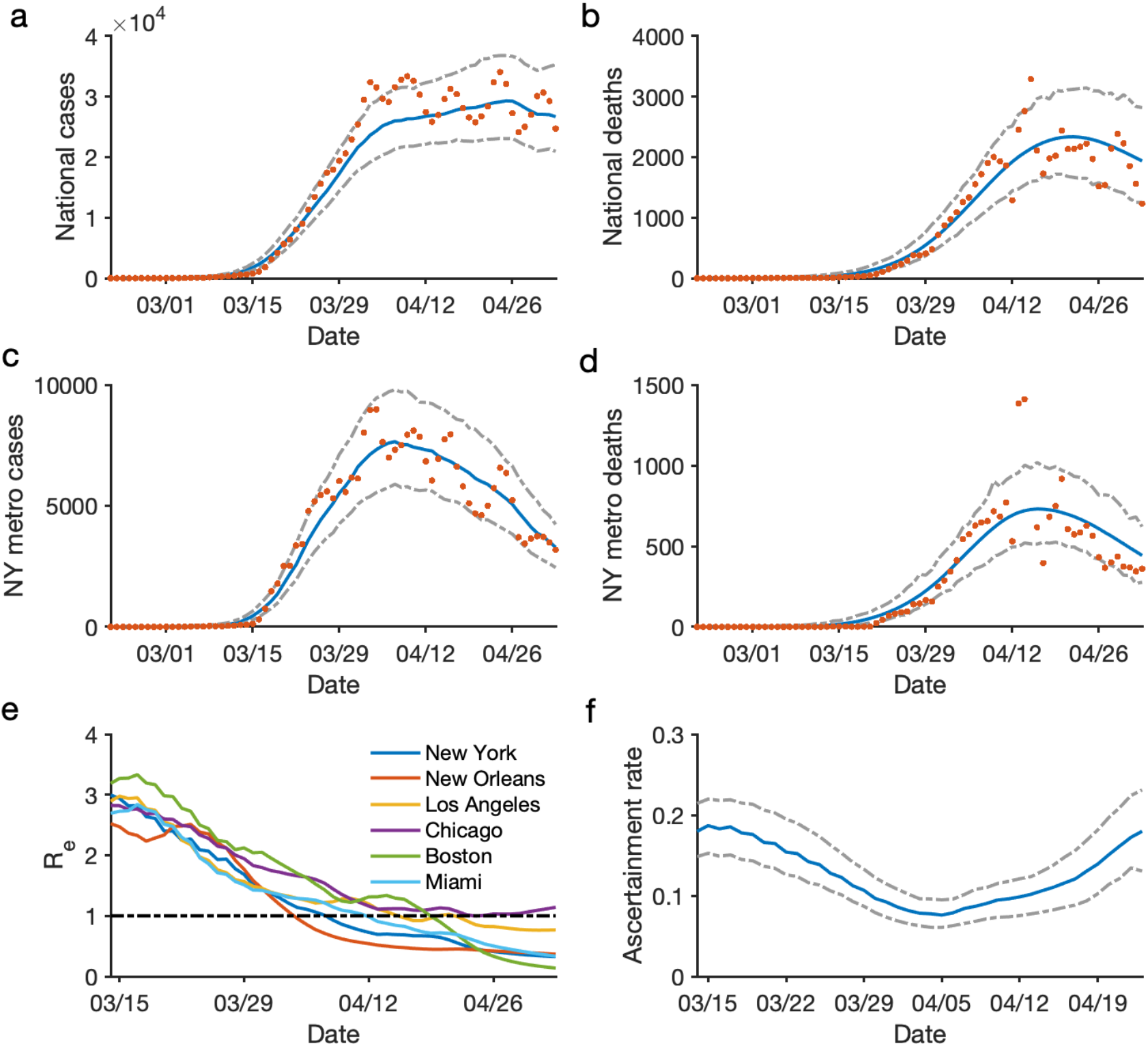
Model fit and parameter inference. Posterior fitting to daily cases and deaths in the US (a-b) and the New York metropolitan area (c-d). Red dots represent observations. Blue and grey lines are the median estimate and 95% CIs. The estimated effective reproductive number, *R_e_*, in six metropolitan areas are shown in (e). The black dotted line indicates *R_e_* = 1. Panel (f) shows the estimated ascertainment rate over time. The blue line and grey lines are the median estimates and 95% CIs.

The inference results indicate that the NPIs varyingly adopted in the US after March 15 have effectively reduced rates of COVID-19 transmission in the focus metropolitan areas. During the initial growth of a pandemic, infections increase exponentially. As a consequence, early intervention and fast response are critical for limiting morbidity and mortality. To quantify the effects of earlier interventions on COVID-19 outcomes in the US, we performed two counterfactual simulations in which the sequence of transmission rates and ascertainment rate inferred for March 15 - May 3, 2020, were shifted back 1 and 2 weeks, i.e. to March 8, 2020 and March 1, 2020, respectively. Specifically, we integrate the transmission model from February 21 to March 8 or March 1, and then apply the daily posterior parameters, i.e., *α* and *β*s, as estimated beginning March 15. The simulations were generated until May 3, 2020. For the last 1-2 weeks without inferred parameters due to the shift in time window, we applied the final parameter estimates of May 3, 2020, the last day of inference.

The counterfactual simulations indicate that had observed control measures been adopted one week earlier, the US would have avoided 645,660 (95% CI: 550,123-753,230) [56.5% (48.1%-65.9%)] confirmed cases and 35,287 (28,473-41,689) [54.0% (43.6%-63.8%)] deaths nationwide as of May 3, 2020 (Fig. 2a-b). In the New York metropolitan area, the epicenter of COVID-19 in the US, 218,397 (195,179-229,394) [83.3% (74.4%-87.4%)] confirmed cases and 18,543 (16,607-19,682) [85.6% (76.2%-90.3%)] deaths would have been avoided if the same sequence of interventions had been applied one week earlier (Fig. 2c-d). A more pronounced control effect would have been achieved had the sequence of control measures occurred two weeks earlier: a reduction of 1,017,544 (956,594-1,066,772) [89.0% (83.6%-93.3%)] cases and 58,332 (54,802-61,297) [89.3% (83.9%-93.9%)] deaths in the US (Fig. 2e-f), and 255,721 (249,316-258,817) [97.5% (95.0%-98.7%)] cases and 21,319 (20,683-21,663) [97.8% (94.8%-99.4%)] deaths in the New York metropolitan area (Fig. 2g-h). These dramatic reductions of morbidity and mortality due to more timely deployment of control measures highlights the critical need for aggressive, early response to the COVID-19 pandemic.

**Figure 2.**
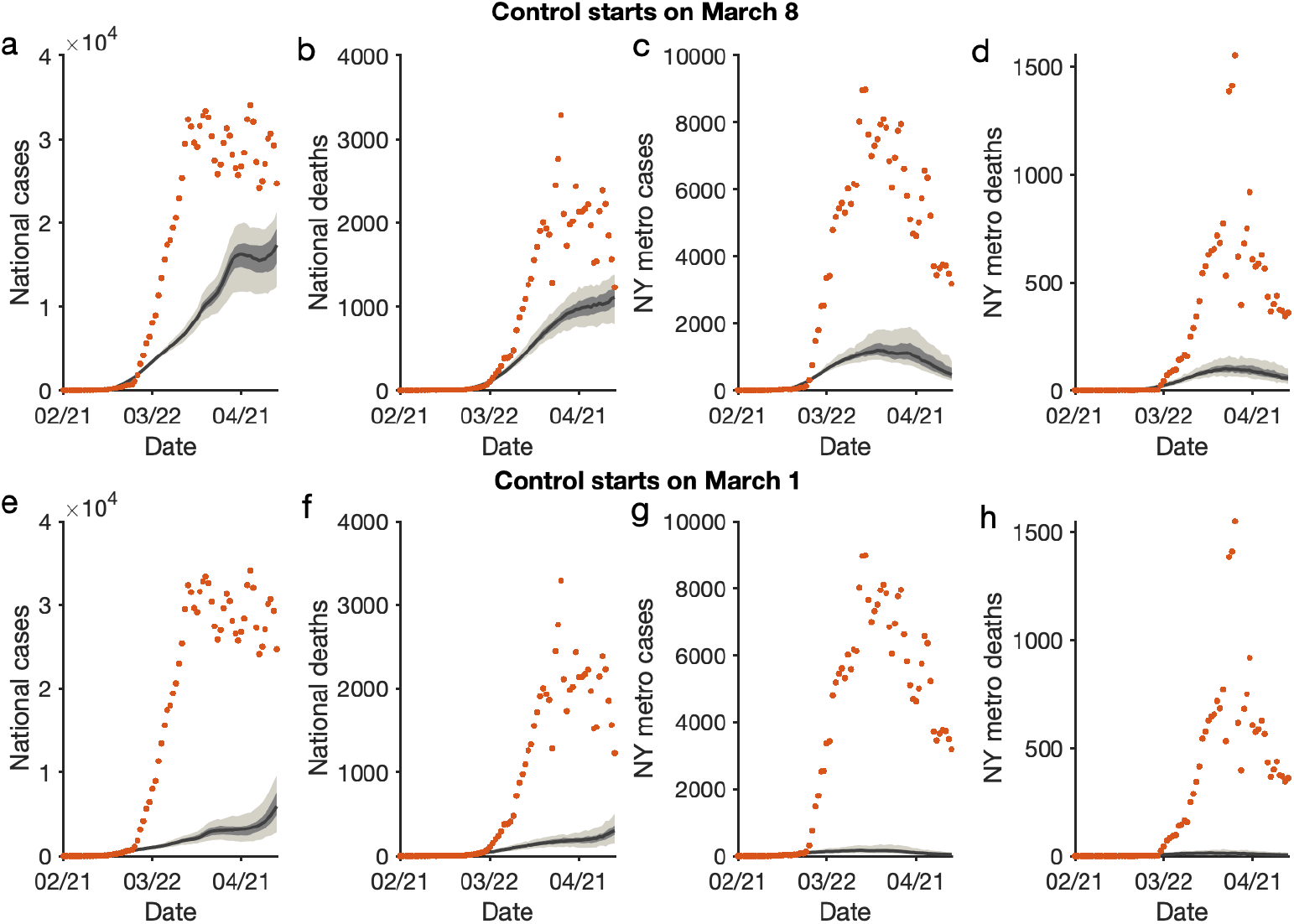
Counterfactual simulations with control interventions beginning in early March – 1 and 2 weeks earlier than implemented. The daily cases and deaths in the US (a,b,e,f) and the New York metropolitan area (c,d,g,h) under early intervention are compared with the observations (red dots). The upper and lower rows present counterfactuals with interventions implemented on March 8 and March 1, respectively. The black lines and surrounding bands show the median estimate, interquartile and 95% CIs.

Now that COVID-19 is established as a global pandemic, rapid response remains essential to avoid large-scale resurgences of infections and deaths in locations with reopening plans. We quantify the effect of response time on the timing and magnitude of rebound in the US through further simulations. Specifically, we assume that control measures are relaxed beginning May 4, 2020 in all US counties, resulting in an increased hypothetical effective reproductive number of *R_e_* = 1.5 in each county. After a response time of 2 or 3 weeks, a reactive 25% weekly reduction of transmission rates, equivalent to the average transmission rate reduction prior to May 4, 2020 (Fig. S5), is imposed in each county and maintained until local weekly case numbers decline.

For both scenarios, a decline of daily confirmed cases continues for almost two weeks after easing of control measures (Fig. 3a-b). This decreasing trend, caused by the NPIs in place prior to May 4, 2020 coupled with the lag between infection acquisition and case confirmation, conveys a false signal that the pandemic is well under control. Unfortunately, due to high remaining population susceptibility, a large resurgence of both cases and deaths follows, peaking in early- and mid-June, despite the resumption of NPI measures 2 or 3 weeks following control relaxation. A one-week further delay to the resumption of control measures results in an average of 32,379 additional deaths nationally by July 1, 2020.

**Figure 3.**
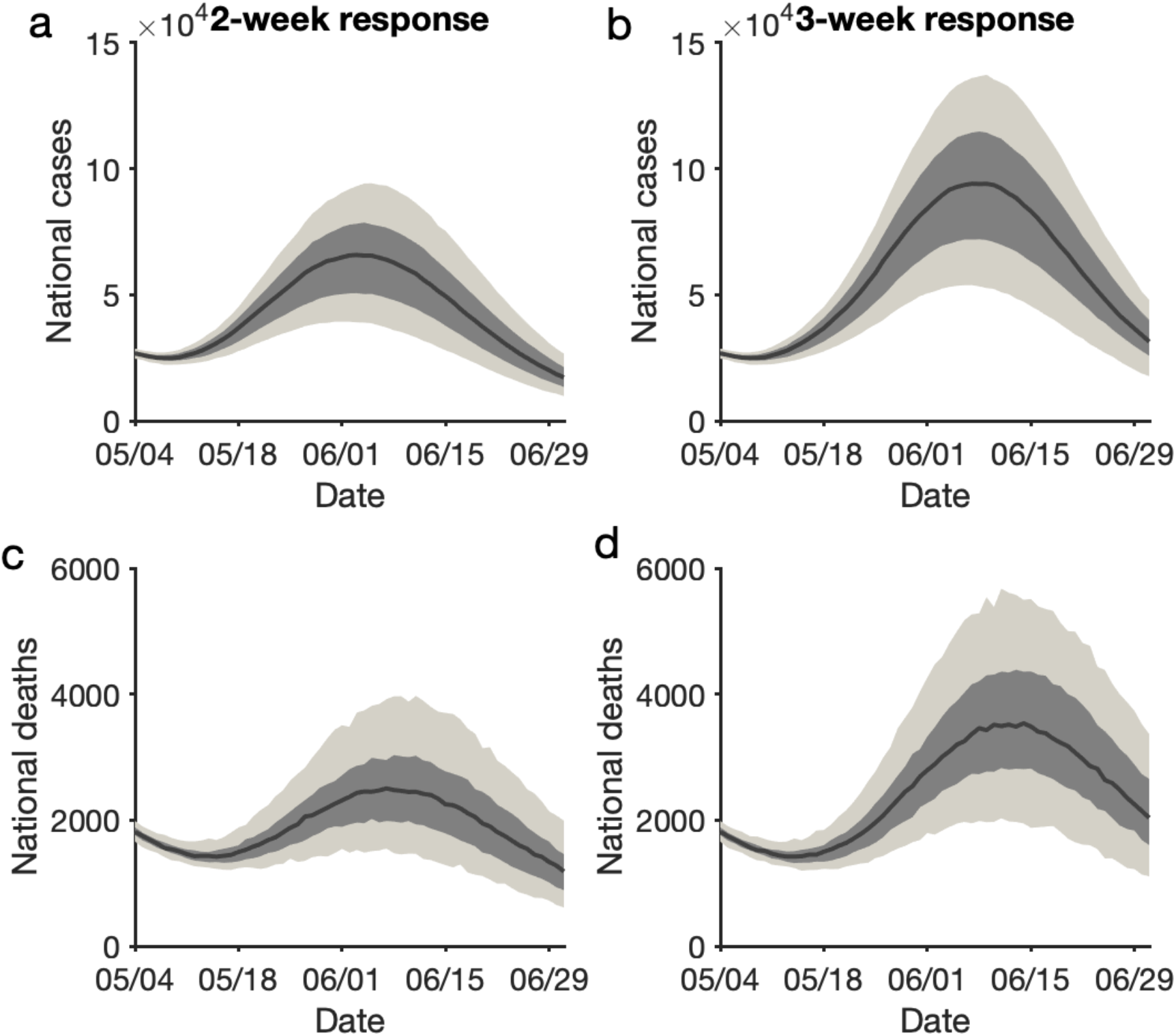
Effects of response time after control measures are relaxed. We assume a control relaxation starting on May 4 in all US counties. After a response time of 2 or 3 weeks, a weekly 25% reduction of the transmission rate is imposed. Daily cases and deaths in the US for a response time of 2 weeks (a,c) and 3 weeks (b,d) are compared. The black lines and bands show the median estimate, interquartile and 95% CIs.

We note these counterfactual experiments are based on idealized hypothetical assumptions. In practice, initiating and implementing interventions earlier during an outbreak is complicated by factors such as general uncertainty, economic concerns, logistics and the administrative decision process. Public compliance with social distancing rules may also lag due to sub-optimal awareness of infection risk. We acknowledge that our counterfactual experiments have simplified these processes; however, we note that by the end of February, 2020, a number of other countries, including South Korea and Italy, were already aggressively responding to the virus^17^. Our findings indicate that had control measures and reductions of *R_e_* in the US been implemented at a similar time, just 1-2 weeks earlier, substantially fewer cases and deaths would have occurred to date.

Our model experiments also indicate that rapid detection of increasing case numbers and fast re-implementation of control measures is needed to control a rebound of outbreaks of COVID-19. In these experiments, we assume the ability to re-implement a 25% weekly reduction of transmission rates nationwide. Due to fatigue in the general public towards NPIs and a consequent reduction in compliance, this assumed reduction may be overly optimistic.

In this study, we have quantified the sensitivity of COVID-19 cases and deaths to the timing of control measures. Our results demonstrate the dramatic impact that earlier interventions could have had on the COVID-19 pandemic in the US. Looking forward, the findings underscore the need for extreme, continued vigilance when control measures are relaxed. While we do not advocate protracted shutdowns and are cognizant of the burdens they impose, it is vital to balance the dual ambitions of renewing economic activity and avoiding a recrudescence.

To date countries such as South Korea, Vietnam, New Zealand and Germany, have shown that such a balance is achievable; the strategies adopted in these countries could be used to guide policies in the US. Specifically, broader testing and contact tracing capacity^18^ are crucial to detect a rebound of COVID-19 before it is well underway^19^. The continued susceptibility to infection of the majority of the US population, including in major metropolitan areas (Fig. S6), can readily support an exponential growth of cases and deaths. Consequently, relaxation of NPI would be more safely effected in localities in which *R_e_* is well below 1, daily confirmed cases are low, and abundant testing and contact tracing are available to aid isolation and quarantine measures.

## Methods

### The metapopulation model

We use a metapopulation SEIR model to simulate the transmission of COVID-19 among 3,142 US counties. In this model, we consider two types of movement: daily work commuting and random movement. Information on county-to-county work commuting is publicly available from the US Census Bureau^12^. We further assume the number of random visitors between two counties is proportional to the average number of commuters between them. As population present in each county is different during daytime and nighttime, we model the transmission dynamics of COVID-19 separately for these two time periods (see supplementary information).

We formulate the transmission as a discrete Markov process during both day and night times. The transmission dynamics are depicted by Eqs. S1-S10 in supplementary information. In these equations, we define *S_ij_*, *E_ij_*, 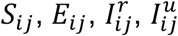 and *N_ij_* as the susceptible, exposed, reported infected, unreported infected and total population in the subpopulation commuting from county *j* to county *i (i ← j)*. We also introduce the following model parameters: *β* is the transmission rate of reported infections; *μ* is the relative transmissibility of unreported infections; *Z* is the average latency period (from infection to contagiousness); *D* is the average duration of contagiousness; *α* is the fraction of documented infections; *θ* is a multiplicative factor adjusting random movement. We integrate Eqs. S1-S10 using a Poisson process to represent the stochasticity of the transmission process.

The transmission model generates daily confirmed cases and deaths for each county. To map infections to deaths, we used an age-stratified infection fatality rate (IFR)^20^ and computed the IFR for each county as a weighted average using demographic information on age structure. To account for reporting delays, we mapped simulated documented infections to confirmed cases using a separate observational delay model. In this delay model, we account for the time interval between a person transitioning from latent to contagious (i.e. 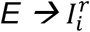) and observational confirmation of that individual infection. To estimate this delay period, *T_d_*, we examined line-list data from early-confirmed cases in China^21^. Prior to January 23, 2020, the time-to-event distribution of the interval (in days) from symptom onset to confirmation is well fit by a Gamma distribution *(a* = 1.85, *b* = 3.57). Consequently, we adopted a Gamma distribution to model *T_d_*, but tested longer mean periods *(ab)*, as symptom onset often lags the onset of contagiousness, using US data prior to March 13, 2020^22^. Our analysis indicates that an average delay of 9 days supports better fit to the daily incidence data. As a result, we adopted *T_d_* = 9 days *(a* = 1.85, *b* = 4.86) in this study. Based on the daily incidence and death data in the US, the national death curve has a 7-day lag compared with the incidence curve (Fig. S7). As a result, we used a gamma distribution with a mean of 16 days (*a* = 1.85, *b* = 8.65) to represent the delay between a person transitioning from latent to contagious and death.

In order to represent variability in transmission rates through space and time, we introduced separate estimates for *β* in the 311 US counties with cumulative cases over 400 as of May 3, 2020. The remaining counties were classified into 16 groups (evenly distributed into a 4 by 4 grouping based on cumulative cases and population density), for which separate transmission rates were defined. In total, 327 transmission rates (*β_i_*) were introduced in the transmission model. Using the next generation matrix approach, we derived the local basic reproductive number, *R*_0_ *= βD[α +* (1 − *α*)*μ*]^16^.

### Data

We used the 2011-2015 5-Year ACS Commuting Flows data from US census survey to prescribe the inter-county movement in the transmission model prior to March 15, 2020, before broad control measures were announced. The county-to-county commuting data is publicly available from the US Census Bureau^12^. We visualize the inter-county commuting in Fig. S1. After March 15, the census survey data are no longer representative due to changes of mobility behavior in response to control measures. Therefore, after March 15, 2020, we use estimates of the reduction of inter-county visitors to points of interest (POI) (e.g., restaurants, stores, etc.)^23^ to inform the decline of inter-county movement on a county-by-county basis. For instance, if the number of inter-county visitors was reduced by 30% in a county on a given day relative to baseline estimates on March 1, 2020, the size of subpopulations traveling to this county would be reduced by 30% accordingly. This real-time mobility data are available between March 1, 2020 and May 2, 2020. For dates beyond May 2, 2020, we maintained the last known level of inter-county movement. We present the evolution of the inter-county mobility index (i.e., the remaining fraction of inter-county visitor numbers relative to the baseline) in the six examined metropolitan areas from March 1 to April 18 in Fig. S2.

County-level daily confirmed cases and deaths were compiled by USAFACTS^11^. Daily cases and deaths in the six metropolitan areas are shown in Figs. S3-S4. At the national scale, we observed a 7-day lag between the reported case and death counts (Fig. S7).

### Model calibration

To derive an estimate of model parameters, we calibrated the transmission model against county-level incidence and death data reported from February 21, 2020 through May 3, 2020. Specifically, we estimated model parameters using a sequential data assimilation technique. The metapopulation model is a high-dimensional system with 60,232 subpopulations. We therefore applied an efficient data assimilation algorithm - the Ensemble Adjustment Kalman Filter (EAKF)^15^, which is applicable to high dimensional model structures, to infer model parameters. The EAKF has been successfully used to infer parameters for seasonal and pandemic influenza^24^ as well as other infectious diseases^25-27^.

To improve the identifiability of this high-dimensional model, we further reduced the number of unknown parameters by fixing disease-related parameters (*Z, D* and *μ*) and the mobility factor (*θ*). These parameters were estimated using the posterior distributions inferred from case data through March 13, 2020^22^. Specifically, we randomly drew these parameters from the posterior ensemble members: *Z* = 3.59 (95% CI: 3.28-3.99), *D =* 3.56 (3.21-3.83), *μ* = 0.64 (0.56-0.70), and *θ =* 0.15 (0.12-0.17).

From February 21, 2020 through May 3, 2020, we performed EAKF inference each day using both case and death data to estimate the ascertainment rate *α* and transmission rates *β_i_*. The prior for the ascertainment rate was drawn from a distribution with a median value *α* = 0.080 (95% CI: 0.069-0.093), estimated in a previous study^22^. The prior transmission rates were scaled based on local population density using the following relation: 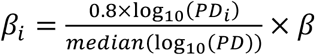. Here *PD_i_* is the population density in county *i*, *median(*log_10_(*PD*)) is the median value of log-transformed population density among all counties, and *β* is the transmission rate estimated before March 13, 2020 *(β* = 0.95, 95% CI: 0.84-1.06)^22^. For *β* shared by multiple counties, population density *PD_i_* is averaged over those counties. To account for reporting delays of confirmed cases and deaths, at each daily model update, we integrated the model forward for 16 days using the prior model state, and used incidence number 9 days ahead and death number 16 days ahead to constrain current model variables and parameters. Given the large number of parameters in the model, the inference system may not be fully identifiable. To alleviate this issue, we imposed a ±30% limit on the daily change of parameters *α* and *β_i_*. This smoothing constraint is reasonable considering the continuity of human’s behavioral change.

In total, we performed 40 independent inference runs. The inference results reported in Fig. 1 were obtained from all posterior ensemble members. Implementation details and system initialization are reported in the supplementary information.

### Metropolitan areas

In this study, we focus on the transmission dynamics in metropolitan areas with dense populations and abundant observations. In particular, we presented parameter estimates for in six metropolitan areas: New York, New Orleans, Los Angeles, Chicago, Boston and Miami. Lists of counties in these metropolitan areas are provided in the supplementary information.

## Data Availability

All input data are publicly available. The counterfactual simulation outcomes for daily cases and deaths in 3142 US counties, as well as the estimated effective reproductive number for the 311 counties with over 400 cumulative cases as of May 3 2020, are available at https://github.com/shaman-lab/Counterfactual.

## Acknowledgements

We thank SafeGraph for sharing the human mobility data. This study was supported by funding from the National Institutes of Health (GM110748) and the National Science Foundation (DMS-2027369), as well as a gift from the Morris-Singer Foundation. The funders had no role in the design, data collection and analysis, decision to publish, or preparation of the manuscript.

## Author contributions

SP and JS designed the study. SP and SK performed the analysis. All authors wrote and reviewed the manuscript.

## Competing interests

JS and Columbia University disclose partial ownership of SK Analytics. JS discloses consulting for BNI.

## Data availability

The counterfactual simulation outcomes for daily cases and deaths in 3142 US counties, as well as the estimated effective reproductive number for the 311 counties with over 400 cumulative cases as of May 3 2020, are available at https://github.com/shaman-lab/Counterfactual.

## Supplementary Information

### Transmission model

We formulate the transmission as a discrete Markov process during both day and night. Daytime transmission lasts for *dt*_1_ days and the nighttime transmission *dt*_2_ days (*dt_1_ + dt*_2_ = 1). Here, we assume daytime transmission lasts for 8 hours and nighttime transmission lasts for 16 hours, i.e., *dt*_1_ = 1/3 day and *dt*_2_ = 2/3 day. The transmission dynamics are depicted by the following equations.

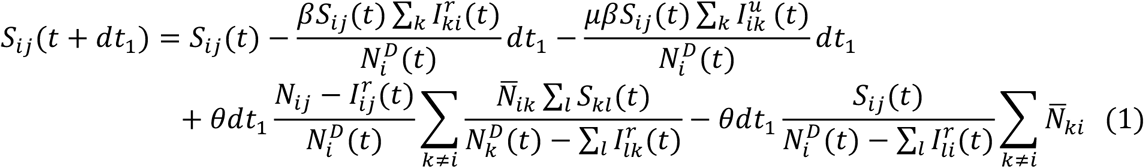

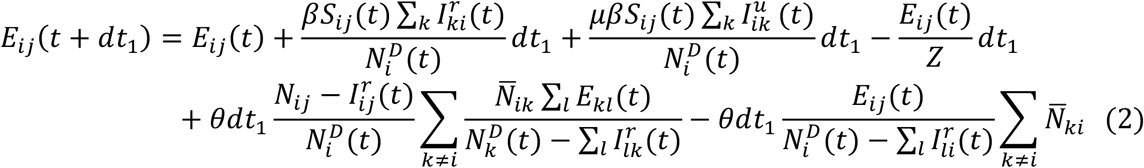

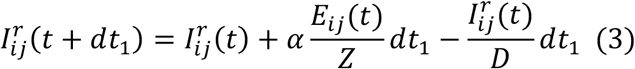

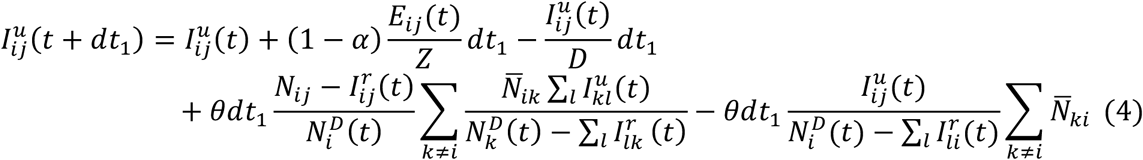

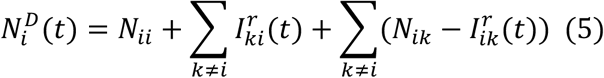

Daytime transmission:

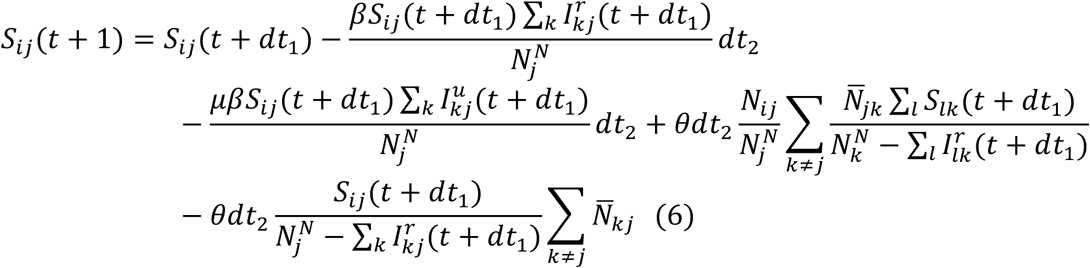

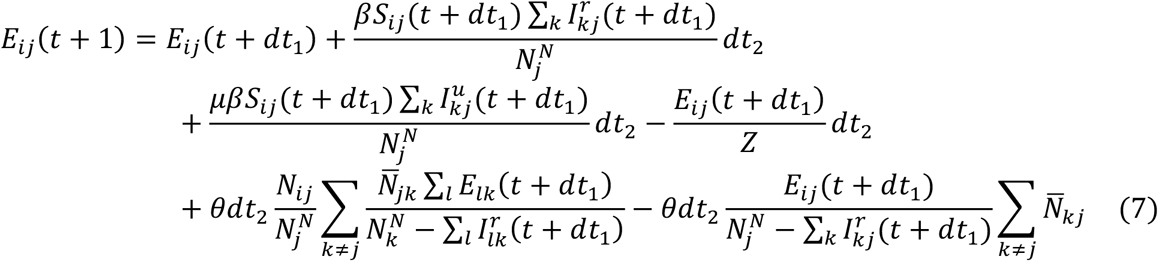

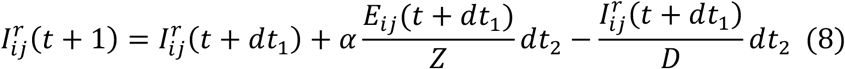

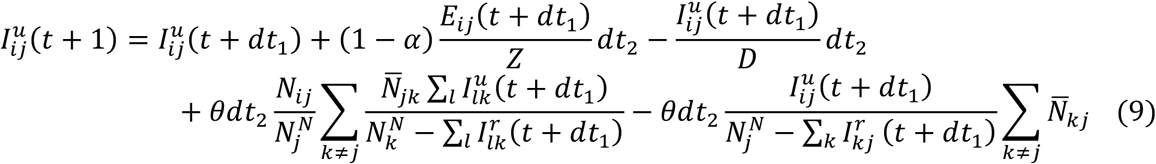

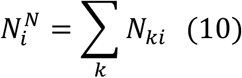

Nighttime transmission:

Here, *S_ij_*, *E_ij_*, 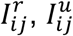 and are the susceptible, exposed, reported infected, unreported infected and total populations in the subpopulation commuting from county *j* to county *i* (*i ← j*); *β* is the transmission rate of reported infections; *μ* is the relative transmissibility of unreported infections; *Z* is the average latency period (from infection to contagiousness); *D* is the average duration of contagiousness; *α* is the fraction of documented infections; *θ* is a multiplicative factor adjusting random movement; 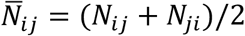 is the average number of commuters between counties *i* and *j;* and 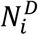 and 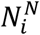 are the daytime and nighttime populations of county *i*.

### The Ensemble Adjustment Kalman Filter

Originally developed for use in weather prediction, the ensemble adjustment Kalman filter (EAKF) assumes a Gaussian distribution of both the prior and likelihood and adjusts the prior distribution to a posterior using Bayes’ rule deterministically. To represent the state-space distribution, the EAKF maintains an ensemble of system state vectors acting as samples from the distribution. In particular, the EAKF assumes that both the prior distribution and likelihood are Gaussian, and thus can be fully characterized by their first two moments (mean and variance). The update scheme for ensemble members is computed using Bayes’ rule (posterior ∝ prior × likelihood) via the convolution of the two Gaussian distributions. For observed state variables, the posterior of the *i*th ensemble member is updated through

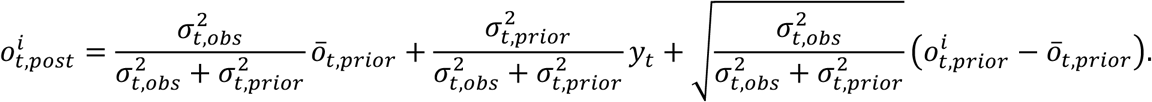

Here 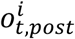 and 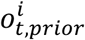 are the posterior and prior of the observed variable (i.e., daily confirmed case or death in each county) for the *i*th ensemble member at time *t; ō_t,prior_* is the mean of the prior observed variable; 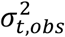 and 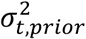 are the variances of the observation and the prior observed variable; and *y_t_* is the observation at time *t*. Unobserved variables and parameters are updated through their covariability with the observed variable, which can be computed directly from the ensemble. In particular, the *i*th ensemble member of unobserved variable or parameter *x^i^* is updated by

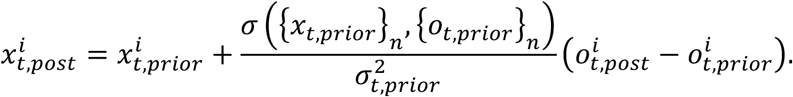

Here 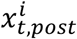 and 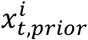 are the posterior and prior of the unobserved variable or parameter for the *i*th ensemble member at time *t*; and 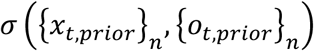 is the covariance between the prior of the unobserved variable or parameter 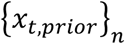 and the prior of the observed variable 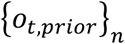 at time *t*. In the EAKF, variables and parameters are updated deterministically such that the higher moments of the prior distribution are preserved in the posterior.

To account for the reporting delay of confirmed case and death, we modified the original EAKF implementation by adjusting model states using observations in the near future, when the effects of parameter change are manifested in observations. Specifically, for data assimilation at day *t*, we ran the transmission model forward to day *t + 16* using prior model state, and used the confirmed case at day *t +* 9 and death at day *t +* 16 to update model variables and parameters at day *t*. This look-ahead data assimilation considered an average delay of 9 days for infection confirmation and an average delay of 16 days for death reporting.

In the EAKF, we assume a heuristic form of observation error variance (OEV) 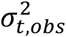. For confirmed cases, we used 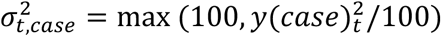, where *y*(*case*)*_t_* is the number of new confirmed cases averaged over day *t — 6* to day *t;* for death, we used 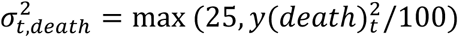, where *y*(*death*)*_t_* is the number of deaths averaged over day *t – 6* to day *t*. Similar forms of OEV have been successfully used for inference and forecasting for a range of infectious diseases. In this study, this OEV setting yields satisfactory fitting.

### System initialization

The prior ranges of model parameters *α* and *β* were set as *α ∈* [0.03,0.25] and *β ∈* [0.01,2.5]. To initialize the model, we seeded exposed individuals (*E*) and unreported infections (*I^u^*) in counties with at least one confirmed case before March 14, 2020. Unlike the situation in China, where the outbreak originated from a single city, importation to multiple locations in the US probably initiated community transmission. To reflect this potential ongoing community transmission before the reporting of the first local infection, for each county with confirmed cases before March 14, we randomly drew *E* and *I^u^* from uniform distributions [0,18*C*] and [0,20*C*] 9 days prior to the reporting date (*T*_0_) of the first case. Here *C* is the total number of reported cases between day *T*_0_ and *T*_0_ *+* 4.

The rationale for this seeding strategy is as follows. If an average reporting delay of 9 days is assumed, we can estimate *I^r^* on day *T*_0_ – 9 as 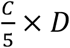, where 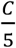 is the average number of daily cases during the first five days with reported cases (*T*_0_ to *T*_0_ *+* 4). If we use the upper bound of 5 days for D, *I^r^* is estimated as *C*, which is also an upper bound. We assume the mean *I^u^* on day *T*_0_ – 9 is 9*C*, implying a reporting rate of 1/10=10%. Drawing *I^u^* from [0,18*C*] leads to a broader prior range of the reporting rate. As both *I^r^* and *I^u^* were evolved from the exposed population *E*, we draw *E* from the range [0,20*C* This crude calculation provides a seeding range for US counties. During inference, this seeding can be adjusted up or down by the filter. The posterior model fittings capture observed outcomes well.

### Counties in six metropolitan areas

We examined counties in six metropolitan areas with cumulative cases over 400 as of May 3, 2020. These counties are:

1. New York: Kings County NY, Queens County NY, New York County NY, Bronx County NY, Richmond County NY, Westchester County NY, Bergen County NJ, Hudson County NJ, Passaic County NJ, Putnam County NY, Rockland County NY
2. New Orleans: Jefferson Parish LA, Orleans Parish LA, St. John the Baptist Parish LA, St. Tammany Parish LA
3. Los Angeles: Los Angeles County CA, Orange County CA
4. Chicago: Cook County IL, DuPage County IL, Kane County IL, McHenry County IL, Will County IL
5. Boston: Norfolk County MA, Plymouth County MA, Suffolk County MA
6. Miami: Miami-Dade County FL, Broward County FL, Palm Beach County FL

**Figure S1.**
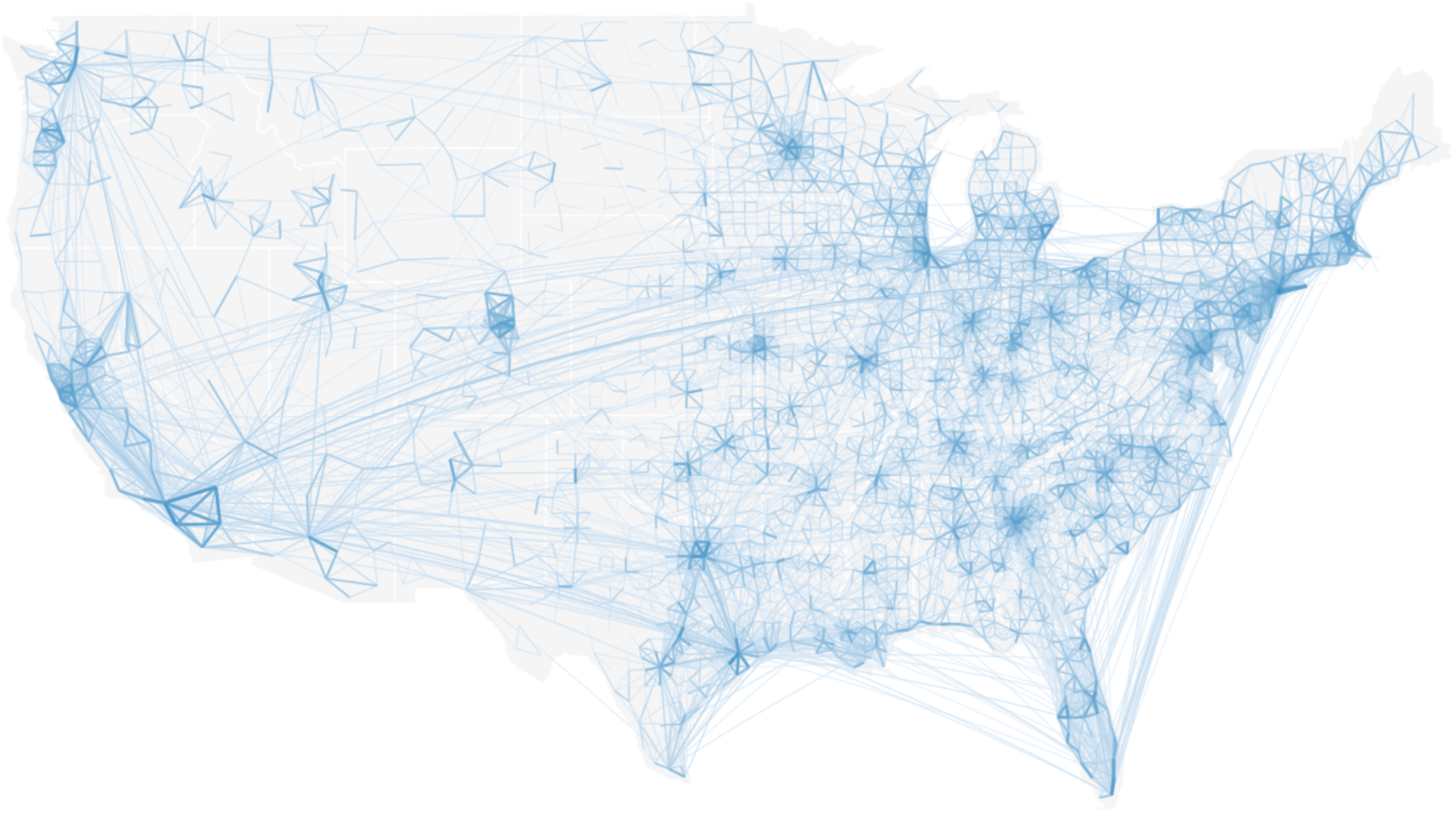
Visualization of inter-county commuting data from US census survey. Line thickness represents the intensity of human movement.

**Figure S2.**
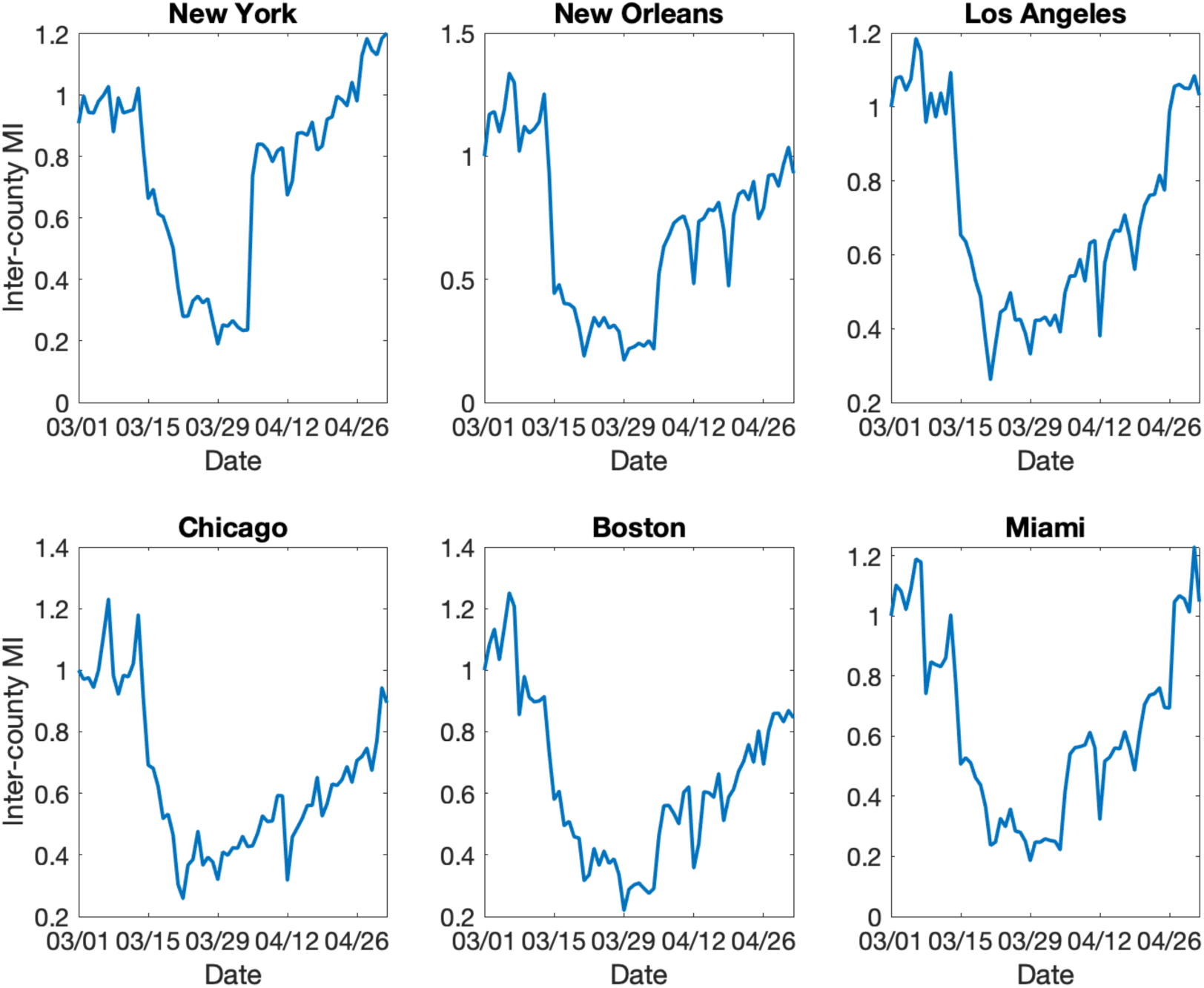
Daily change of inter-county human movement in six metropolitan areas. The inter-county mobility index (MI) is defined as the fraction of inter-county visitors relative to the baseline on March 1, 2020.

**Figure S3.**
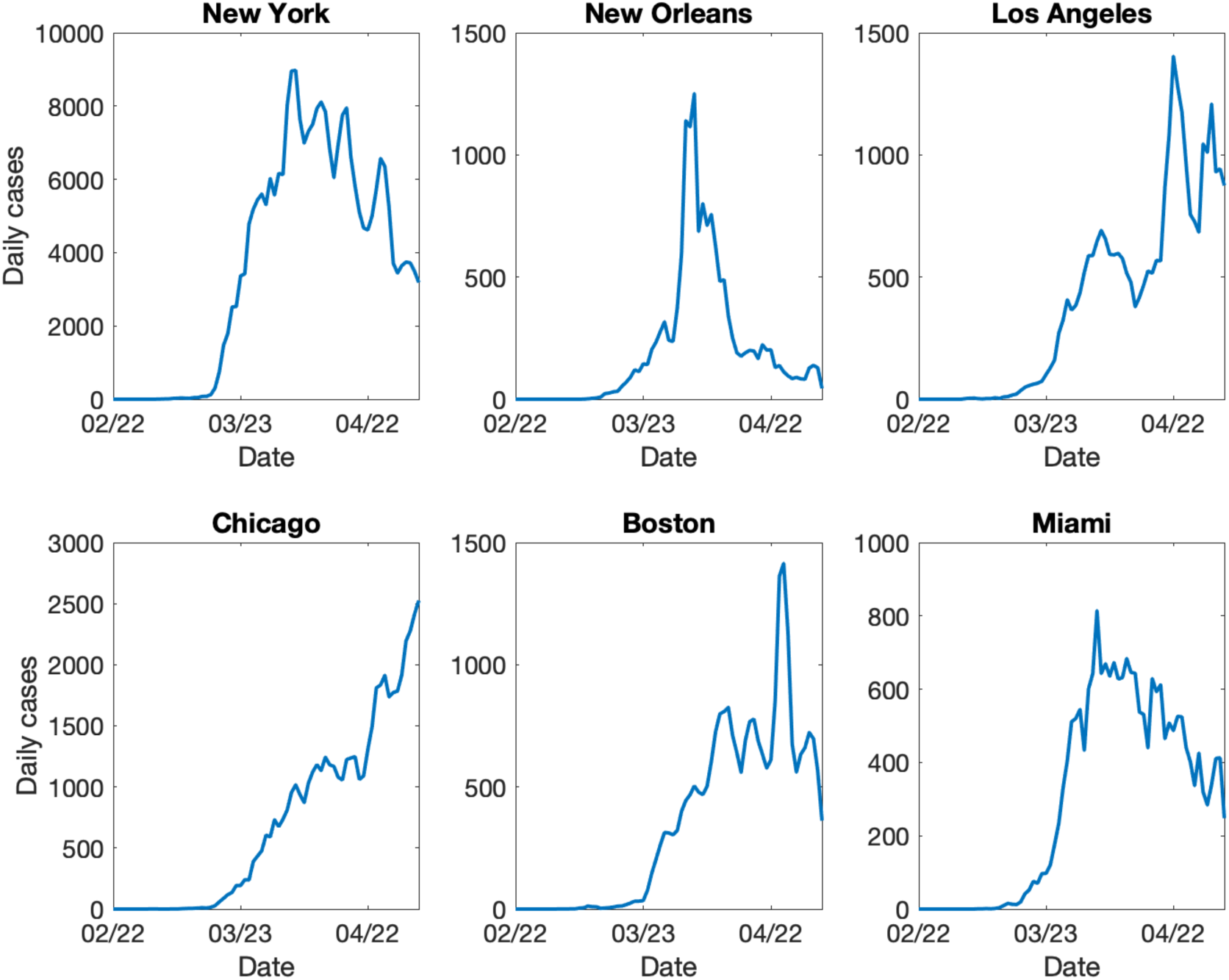
Daily confirmed cases in six metropolitan areas as of May 3, 2020.

**Figure S4.**
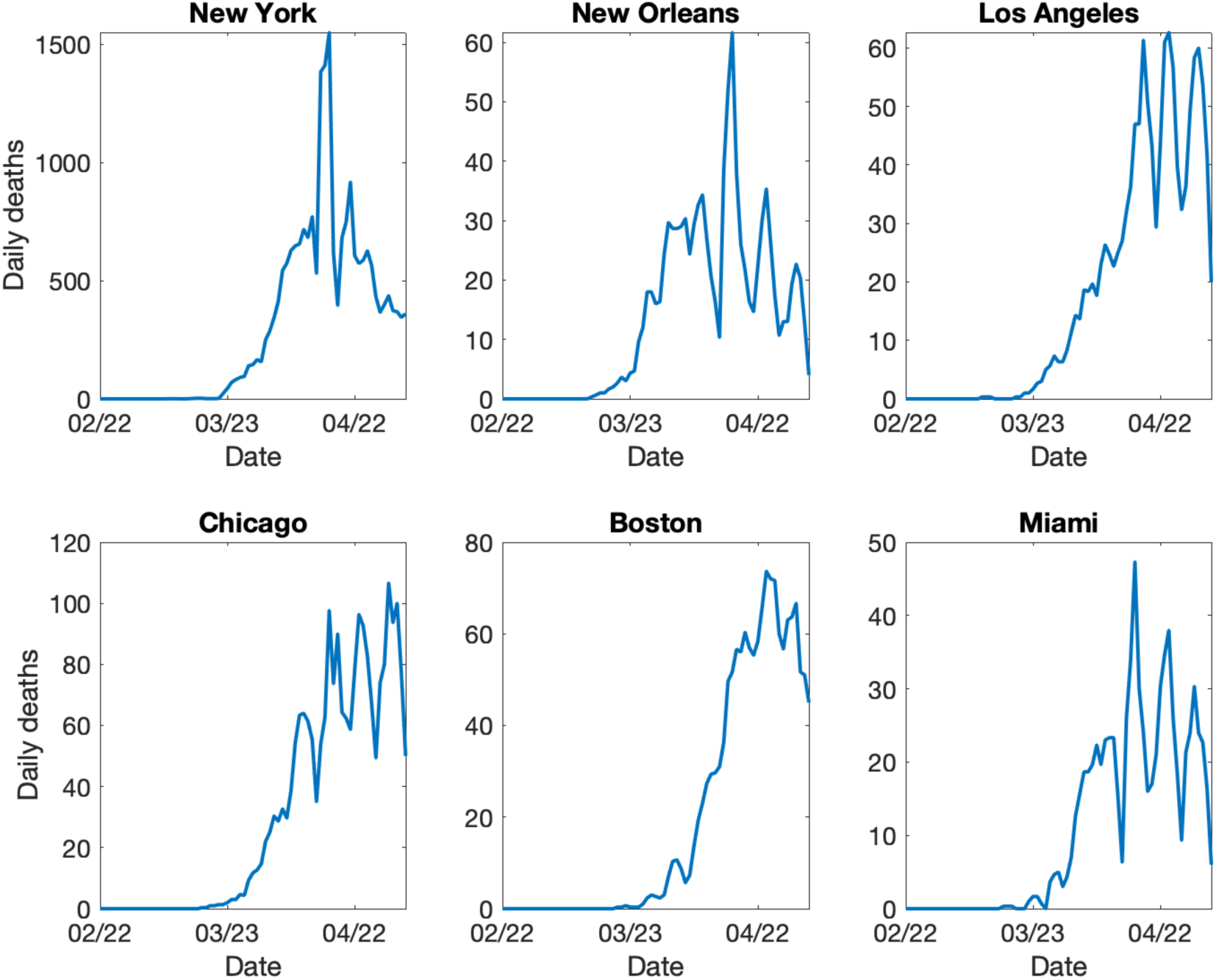
Daily reported deaths in six metropolitan areas as of May 3, 2020.

**Figure S5.**
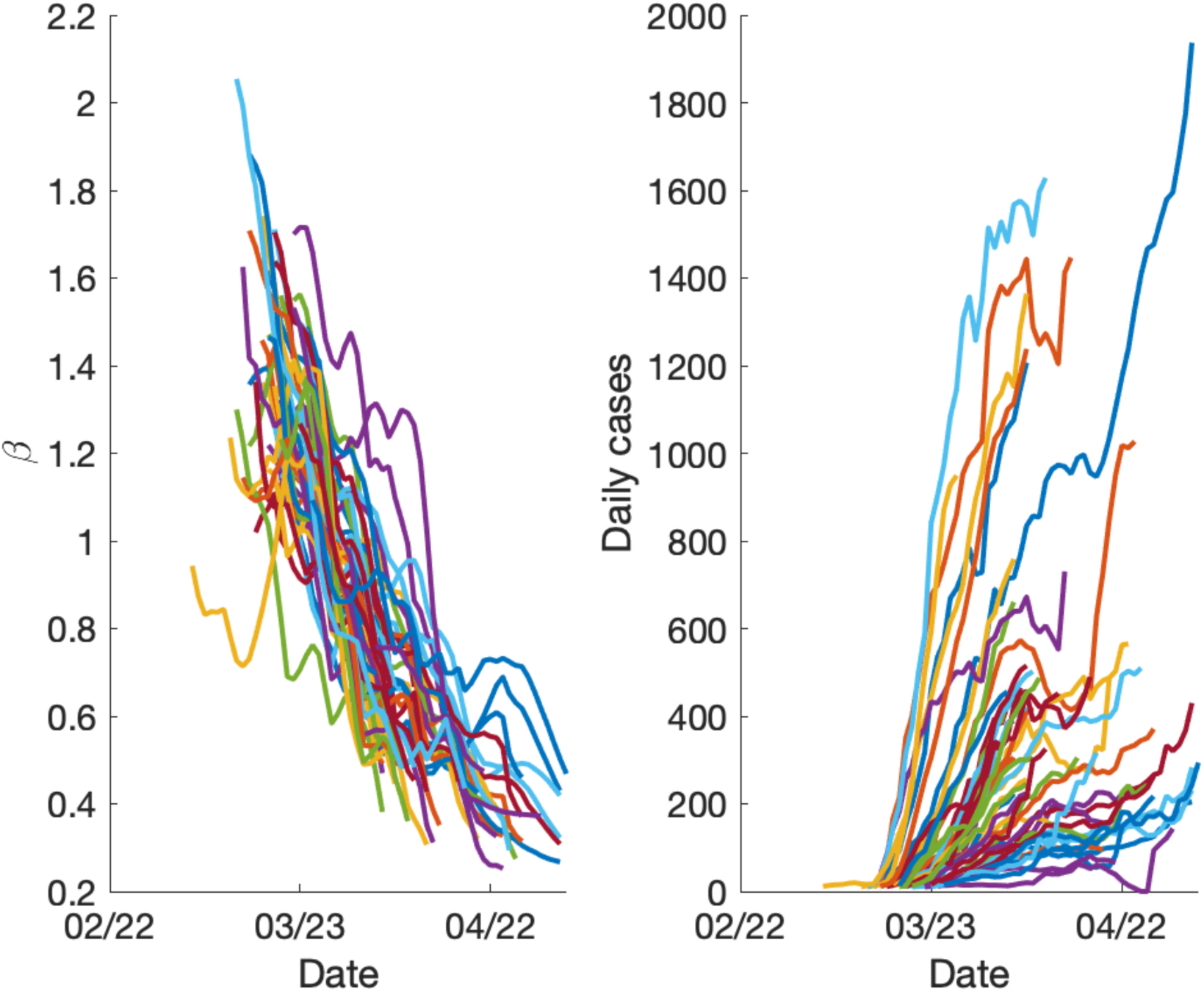
The reduction of transmission rates in counties with increasing confirmed cases. We selected segments of increasing cases for counties with a maximum daily case level above 200 (right panel), and inspected the estimated transmission rates in those counties during the same period (left panel). The average weekly reduction of transmission rate is 25% in those counties with increasing confirmed cases.

**Figure S6.**
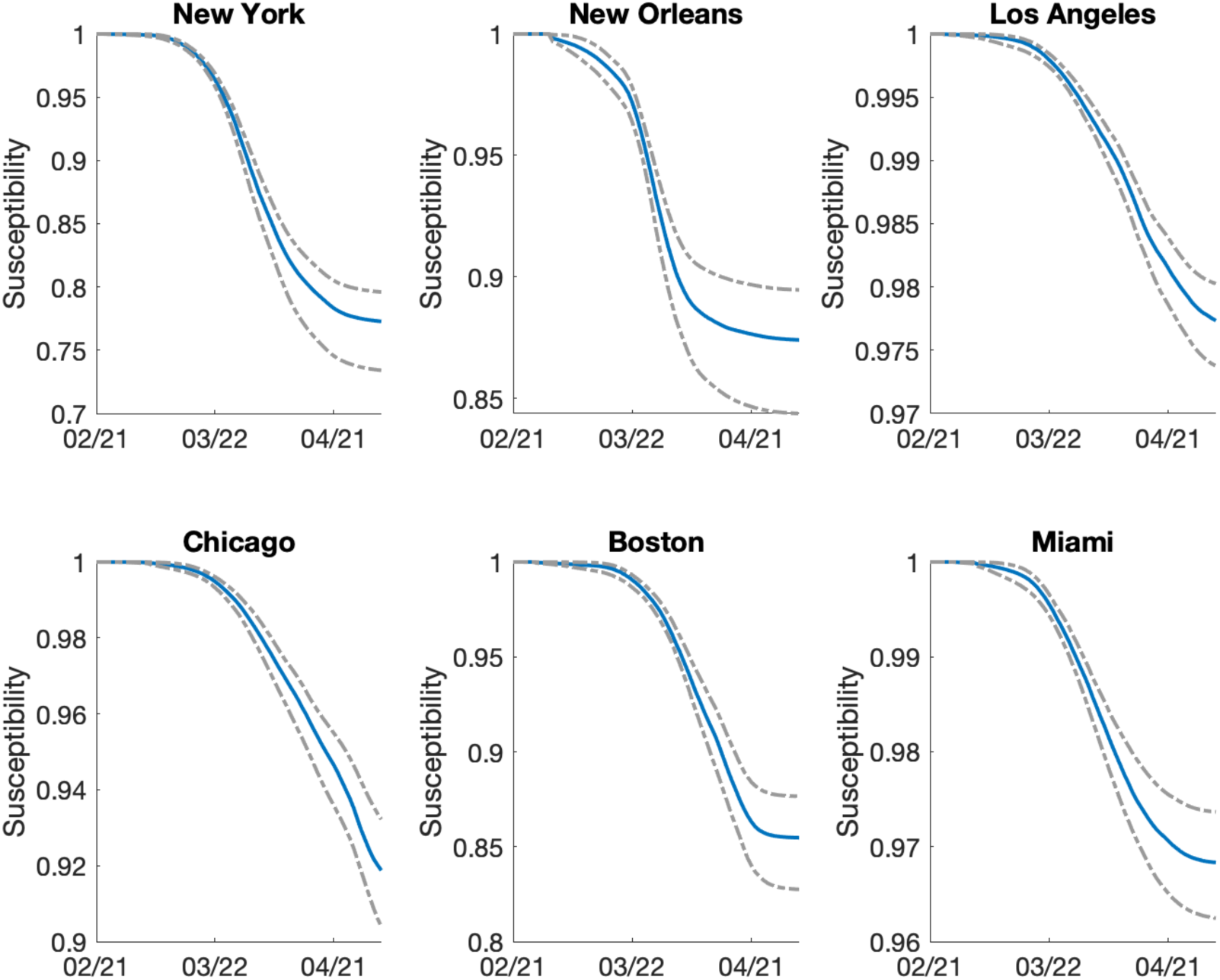
The estimated fraction of susceptible population in six metropolitan areas. Blue line is the median estimate and grey dotted lines are 95% CIs.

**Figure S7.**
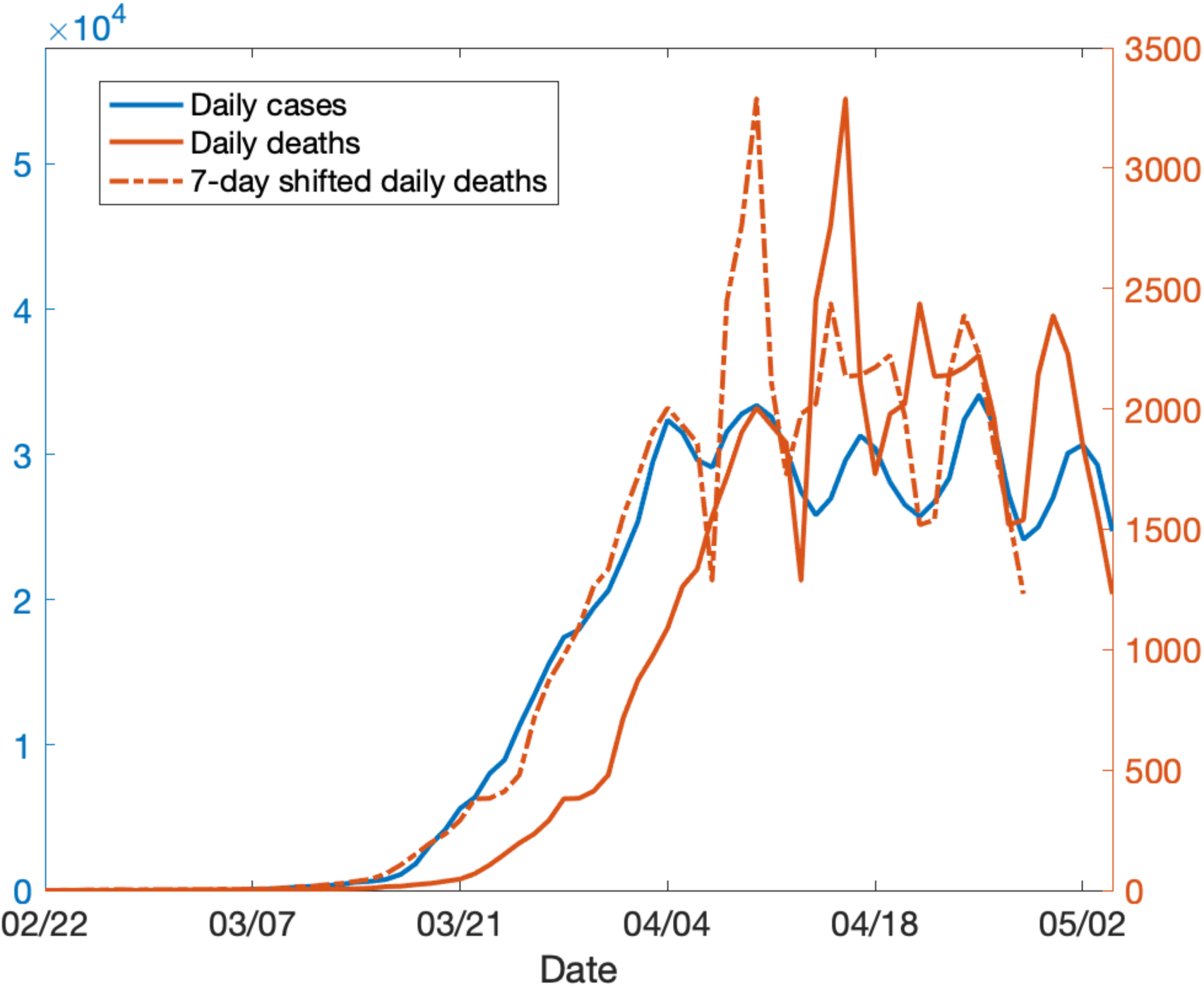
National daily confirmed cases and deaths. The dotted red line is the death time series shifted 7 days backwards. A 7-day delay between the curves of confirmed cases and deaths is observed.

## Notes

### Author Declarations

All data are anonymized or are publicly available.

